# Clinical and Computational Speech Measures are Associated with Social Cognition in Schizophrenia Spectrum Disorders

**DOI:** 10.1101/2022.03.18.22272633

**Authors:** Sunny X. Tang, Yan Cong, Amir H. Nikzad, Aarush Mehta, Sunghye Cho, Katrin Hänsel, Sarah Berretta, John Kane, Anil K Malhotra

**Affiliations:** Zucker Hillside Hospital, Department of Psychiatry, Feinstein Institutes for Medical Research, 75-59 263rd St., Glen Oaks, NY 11004; University of Pennsylvania, Linguistic Data Consortium, 3600 Market St., Suite 810, Philadelphia, PA 19104; Yale University, Department of Laboratory Medicine, 195 Church Street, New Haven, CT 06510

**Keywords:** schizophrenia, language, disorganization, natural language processing, social cognition, computational

## Abstract

In this study, we compared three domains of social cognition (emotion processing, mentalization, and attribution bias) to clinical and computational language measures in 63 participants with schizophrenia spectrum disorders. Based on the active inference model for discourse, we hypothesized that emotion processing and mentalization, but not attribution bias, would be related to language disturbances. Clinical ratings for speech disturbance assessed disorganized and underproductive dimensions. Computational features included speech graph metrics, use of modal verbs, use of first-person pronouns, cosine similarity of adjacent utterances, and measures of sentiment; these were represented by four principal components characterizing content-rich speech, insular speech, local coherence, and affirmative speech. We found that higher clinical ratings for disorganized speech predicted greater impairments in both emotion processing and mentalization, and that these relationships remained significant when accounting for demographic variables, overall psychosis symptoms, and verbal ability. Similarly, computational features reflecting insular speech also consistently predicted greater impairment in emotion processing. There were notable trends for underproductive speech and decreased content-rich speech predicting mentalization ability. Exploratory longitudinal analyses in a small subset of participants (n=17) found that improvements in both emotion processing and mentalization were predicted by improvements in disorganized speech. Attribution bias did not demonstrate strong relationships with language measures. Altogether, our findings are consistent with the active inference model of discourse and suggest greater emphasis on treatments that target social cognitive and language systems.

## 1. Introduction

Social cognitive impairments and speech disturbances are prominent characteristics of schizophrenia spectrum disorders (SSD). Both are linked to canonical SSD neural pathways and brain systems (Green et al., 2015; Kircher et al., 2018); clinically, both are related to poor outcomes and decreased functioning (Bowie et al., 2011; Couture et al., 2006; Schmidt et al., 2011; Tan et al., 2014). Several have suggested that social cognitive impairments and speech disturbances may share these characteristics not by coincidence - but rather, they are causally related to one another and to the pathogenesis of psychosis in general (Palaniyappan and Venkatasubramanian, 2022; Wible, 2012).

Social cognition can be understood in terms of a lower-level simulation factor, which encompasses first-order representations of others’ mental states, and a higher-level mentalizing factor, which formulates complex mental state attributions (Oliver et al., 2019). Alternatively, four domains can be identified, including emotion processing, mentalization, attribution bias, and social perception (Green et al., 2008; Pinkham et al., 2018). Whichever framework is used, consistent impairments are found in SSD for all domains and social cognitive factors (Green et al., 2012; Oliver et al., 2020). Moreover, these impairments are also present among individuals at clinical and genetic risk for psychosis (Green et al., 2012; Kohler et al., 2014; Tang et al., 2017).

Speech-related disturbances in SSD have previously been equated with thought disorder (Andreasen, 1979). While the parallels between speech and thought are intriguing, in this study, we have focused on speech qua speech. Commonly, speech production disturbances in SSD can be understood as disorganized (roughly equivalent to positive thought disorder and incoherence) or underproductive (roughly equivalent to negative thought disorder, impoverishment, and alogia) (Kircher et al., 2018). As with social cognition, speech disturbances have also been identified among individuals at clinical and genetic risk for psychosis (Corcoran et al., 2018; Morgan et al., 2017; Solot et al., 2020).

The Bayesian active inference model of discourse can be interpreted to imply a direct link between social cognition and speech (Brown and Kuperberg, 2015; Palaniyappan and Venkatasubramanian, 2022; Vasil et al., 2020). Simplistically, this model describes an active feedback process where the speaker monitors internal and, critically, external signals during speech production. When an error is detected, i.e., a mismatch between predicted versus perceived outcomes, the “prior” representation for the intended communication may be updated. For example, if the speaker begins to discuss how “Mary said something interesting,” but perceives confusion on the part of their listener, the speaker may introduce additional information, “you know, the postdoc I work with.” Updating the prior too often or with inaccurate error estimates could then result in apparently disorganized speech. Thus, impairments in emotion processing and mentalization might directly translate to speech disturbance. Other aspects of the model have also been used to explain receptive language impairments and the presence of hallucinations and delusions in psychosis (Brown and Kuperberg, 2015).

Experimental results have largely borne out the relationship between social cognition and speech suggested by the active inference model. Healthy volunteers were found to adjust how they referenced entities based on assumptions about their conversation partners’ mental states (Achim et al., 2017). Similarly, mentalization ability among people with SSD was associated with the degree to which they aligned their word usage with that of the experimenter (Dwyer et al., 2019). In a meta-analysis of 123 studies, there was a moderate association between clinical ratings related to speech (disorganization, alogia, or thought disorder) and mentalization (r = -0.35) as well as emotion recognition (r = -0.33), but smaller effect sizes for social perception, emotion regulation, and attribution bias (r = 0.1-0.2) (de Sousa et al., 2019). When accounting for non-social neurocognition, emotion processing and mentalization each explained additional variance in speech disturbances (Docherty et al., 2013). Notably, these studies have largely used subjective clinical ratings for speech. In one exception, an aggregated measure of social cognition was found to be modestly associated (r=-0.28) with a computational measure of simplicity (Minor et al., 2019). However, a limited number of features was examined and it remains to be determined how different domains of social cognition relate to objective measures of speech disturbance in SSD.

In this study, we examined the relationships between social cognition and subjective clinical ratings for speech disturbance alongside objective computational features derived from automated analyses. The domains of emotion processing, mentalization, and attribution bias were evaluated. Based on the Bayesian active inference model for discourse, we hypothesized that emotion processing and mentalization would be significantly related to clinical and computational measures of speech disturbance in SSD, even when accounting for variability in overall psychosis symptoms and verbal ability. We further expected that attribution bias would be less directly related to speech disturbance as it does not have an integral function in the active inference model for discourse.

## 2. Methods

### 2.1 Participants and Overall Design

Participants with SSD (N=63) were recruited from inpatient and outpatient facilities at Zucker Hillside Hospital in Queens, New York, and underwent informed consent (Table 1). All procedures were approved by the Institutional Review Board at the Feinstein Institutes for Medical Research. A subset returned for a follow-up assessment after 12 weeks (n=17). The longitudinal subsample was primarily recruited from stable outpatients, so they had milder symptoms and higher social cognitive ability than those who did not return for a follow-up visit.

**Table 1.**
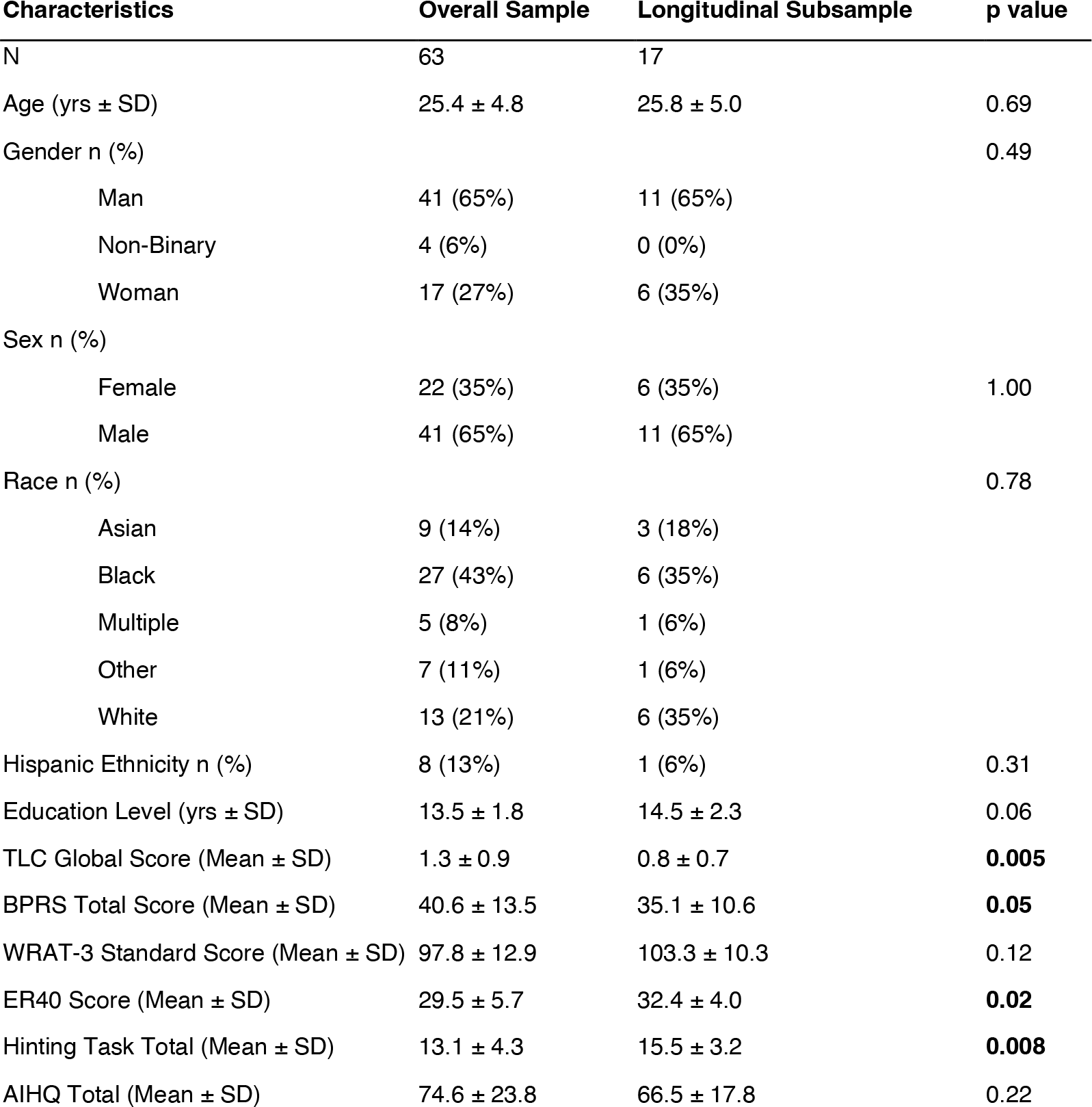
Participant Characteristics. Note: TLC Global Score – derived from the global rating of speech and language disturbance (0-4) based on the Scale for the Assessment of Thought, Language and Communication. A score of 1.3 falls between “mild” and “moderate” TLC disorder; a score of 0.8 falls between “none” and “mild.” Significant differences between the longitudinal subsample and overall sample are bolded. Yrs – years. SD – standard deviation. BPRS – Brief Psychiatric Rating Scale. ER40 – Penn emotion recognition task. AIHQ - Ambiguous Intentions and Hostility Questionnaire

Participants were screened with the psychosis and mood portions of the Structured Interview for the DSM-IV (First and Gibbon, 2004) and met DSM-5 criteria for schizophrenia spectrum disorders (schizophrenia: n=39 schizophrenia; schizoaffective disorder: n=10; unspecified psychotic disorder: n=8; schizophreniform disorder: n=5; delusional disorder: n=1) (American Psychiatric Association, 2013). Dimensional ratings of overall psychosis symptoms were obtained using the Brief Psychiatric Rating Scale (BPRS) (Overall and Gorham, 1962). Verbal ability was assessed with the reading portion of the Wide Range Achievement Test-3 (WRAT-3) (Snelbaker et al., 2001). In addition, all participants provided recorded responses to three picture description tasks and two open-ended narrative prompts (e.g., “Tell me about yourself”). Recordings were made either in person via a standalone recorder (n=15), virtually through video conferencing software (n=19), or via an integrated iPad app developed by Winterlight Labs with in-person research coordinators facilitating the process (n=29). There was a significant interaction between data collection method and ascertainment, as all virtual participants were stable outpatients, all participants through the iPad app were inpatients, and most in person assessments were with inpatients.

### 2.2 Social Cognitive Assessments

Social cognition domains were assessed with validated instruments with good psychometric properties (Buck et al., 2017; Pinkham et al., 2018). Emotion processing was assessed with the Penn Emotion Recognition Task (ER-40), a computerized battery where participants identify 40 images of faces as angry, fearful, sad, happy or no emotion (Kohler et al., 2003; Moore et al., 2015). Mentalization was assessed with the Hinting Task (Corcoran et al., 1995) using revised scoring from the SCOPE study to decrease the ceiling effect (Pinkham et al., 2018). Participants read 10 vignettes about social interactions and then answered questions regarding the characters’ intentions. Attribution bias was assessed with the Ambiguous Intentions and Hostility Questionnaire (AIHQ), where participants rate their perceptions of intentionality, anger, and blame for social vignettes (Combs et al., 2007). Per the psychometric evaluations by Buck et al., we used the overall blame scores from 5 ambiguous and 5 accidental scenarios with higher scores signifying greater attribution of hostility and intentionality (Buck et al., 2017). Table 1 reports raw scores. Because the ER-40 and Hinting Task scores were left-skewed, we transformed and standardized the scores via the Yeo-Johnson method prior to statistical analyses (Yeo and Johnson, 2000). AIHQ scores were normally distributed and z-transformed.

### 2.3 Clinical Language Ratings

Participants were rated on 18 items and a global score using the Scale for the Assessment of Thought Language and Communication (TLC) (Andreasen, 1986). The TLC includes items associated with disorganization / positive thought disorder (e.g., tangentiality, incoherence, derailment), as well as underproductive speech / negative thought disorder (poverty of speech). The TLC items for clanging, word approximations, echolalia, blocking, stilted speech, and self-reference were omitted from the assessment because they were uncommon and interrater reliability was not ideal (ICC < 0.9). Because we wished to study speech disturbance in general, we additionally included the SANS items for latency and decreased vocal inflection. Coordinators underwent an iterative training process and demonstrated excellent interrater reliability for the included items (ICC ≽ 0.9). Ratings were provided for each participant holistically, based on the entire research encounter.

### 2.4 Computational Language Measures

Recordings from the three picture descriptions and two open-ended narratives were transcribed verbatim by human annotators and reviewed for accuracy prior to automated analyses; disfluencies such as filled pauses, incomplete words and repeats were manually tagged during transcription and removed. Utterance boundaries were determined manually based on syntactic completeness and the presence of pauses. Within each utterance, tokens were identified by NLTK word-tokenizer (Loper and Bird, 2002). Each token was tagged for its part-of-speech (POS) and lemmatized using spaCy modules (Honnibal and Johnson, 2015). All computational processing was completed using Python.

We chose features from five computational strategies based on a priori hypotheses that they would be sensitive to differences in social cognitive ability and the active inference process. 1) Network properties of speech graphs quantify the scope and interconnectedness of discourse (Mota et al., 2012). These features have been associated with disorganized and impoverished speech in psychosis (Mota et al., 2017; Palaniyappan et al., 2019). Consistent with the methods described by Mota et al. (2012), we formed speech graphs by connecting lexemes (nodes) based on their sequential relationships (edges). Graph features include density, diameter, average shortest path length (ASPL), largest strongly connected component (LSCC), largest clique, average weighted degree, and average clustering coefficient. Counts of nodes and edges were highly correlated with average degree, so were not included. 2) Modal verbs like “should,” “will,” “can,” and “could” express states of possibility, obligation and intention, which is relevant to mentalization. We quantified the use of modal verbs by counting their occurrence and standardizing by the number of total verb phrases. 3) Previous work found that individuals with SSD were more likely to use first person singular pronouns and less likely to use first person plural pronouns (Tang et al., 2021). We quantified the number of first person singular and plural pronouns and standardized the counts based on total word count from each task. Only the open-ended narrative tasks were used because first person pronouns are not expected consistently on picture description tasks. 4) Local coherence, as measured by the cosine similarity between pairs of adjacent utterances, has been shown to be a predictor of dimensional speech disturbance, SSD diagnosis, and later development of psychosis among youths at clinical high risk (Bedi et al., 2015; Corcoran et al., 2018; Iter et al., 2018; Krell et al., 2021). We computed cosine similarities using mean embeddings from GloVe (Pennington et al., 2014), Word2Vec (Mikolov et al., 2013), and latent semantic analysis (LSA) (Elvevåg et al., 2007; Landauer et al., 1998). 5) Speech sentiment may be related to emotion processing as well as mentalization, and was quantified using standardized measures of valence (i.e., happy vs. sad), arousal (i.e., excited vs. calm), and dominance (i.e., in control vs. controlled) (Warriner et al., 2013).

### 2.5 Statistical Analyses

Statistical analyses were completed in R (R Core Team, 2016). To reduce the dimensionality of the clinical and computational language measures, principal component analyses using the promax rotation were conducted with the *psych* package (Revelle, 2021). Based on visual examination of the scree plots, two components were chosen for the clinical ratings, and four components were chosen for the computational features. Spearman coefficients were used to calculate the correlations among social cognition and language measures. To account for multiple comparisons (3 domains × 2 clinical components or 3 domains × 4 computational components), we adjusted p values for the correlations using the false discovery rate (FDR) method (Benjamini and Hochberg, 1995). To evaluate potentially confounding relationships with data collection method, demographic variables (age, sex, education level), overall psychosis symptoms, and verbal ability, we evaluated multiple linear regressions with these as covariates. All reported *β* coefficients are standardized. Longitudinal relationships between social cognition and language measures were evaluated using linear mixed models from the *nlme* package (Pinheiro et al., 2022). We predicted social cognition with fixed effects of timepoints and language measures, and random effects of participants.

## 3. Results

### 3.1 Principal Components of Speech Measures

Table 2 details principal components and item loadings for clinical ratings and computational language features. The two-component solution for clinical language ratings yielded expected results: (1) Disorganized Speech, and (2) Underproductive Speech. The four-component solution for computational language features yielded the following: (1) Content-Rich Speech, characterized by more extensive unique content and use of modal verbs; (2) Insular Speech, characterized by greater quantities of speech focused around the same interconnected content and use of first person singular pronouns like “I”; (3) Local Coherence, characterized by cosine similarities between mean embeddings of adjacent sentence pairs; and (4) Affirmative Speech, characterized by positive valence, high dominance (“in control”), low arousal (“calm”), and use of first-person plural pronouns like “we”.

**Table 2.** Principal Components of Clinical and Computational Speech Measures. Note: Loadings above an absolute value of 0.3 are shown. TLC – Scale for the Assessment of Thought Language and Communication. SANS – Scale for the Assessment of Negative Symptoms. ASPL – Average shortest path length between any two nodes. LSCC – Number of nodes in the largest strongly connected graph component. LSA – Latent semantic analysis

| A) Clinical Items | C1 -<br>Disorganized | C2 -<br>Underproductive | B) Computational Items | C1 - Content-Rich | C2 -<br>Insular | C3 - Local<br>Coherence | C4 -<br>Affirmative |
| --- | --- | --- | --- | --- | --- | --- | --- |
| TLC1 - Pov. Speech |  | 0.87 | Graph - Avg. Degree |  | 0.88 |  |  |
| TLC2 - Pov. Content Speech | 0.80 |  | Graph - Density | -0.92 |  |  |  |
| TLC3 - Pressure | 0.74 |  | Graph - ASPL | 0.86 |  |  |  |
| TLC4 - Distractible | 0.63 |  | Graph - Clustering Coeff. | -0.43 | 0.66 |  |  |
| TLC5 - Tangentiality | 0.87 |  | Graph - Largest Clique | 0.31 | 0.91 |  |  |
| TLC6 - Derailment | 0.80 |  | Graph - LSCC | 0.82 | 0.42 |  |  |
| TLC7 - Incoherence | 0.50 | 0.36 | Graph - Diameter | 0.90 |  |  |  |
| TLC8 - Illogicality | 0.71 |  | Modal Auxiliary Verbs | 0.46 |  |  |  |
| TLC10 - Neologism | 0.32 |  | Embedding - Glove |  |  | 0.86 |  |
| TLC12 - Circumstantiality | 0.80 | -0.31 | Embedding - LSA |  | 0.47 | 0.63 |  |
| TLC13 - Loss of Goal | 0.68 | 0.37 | Embedding - Word2Vec |  |  | 0.88 |  |
| TLC14 - Perseveration | 0.79 |  | Pronouns - 1st Per. Singular | -0.42 | 0.51 |  |  |
| SANS6 - Dec. Vocal Inflection |  | 0.69 | Pronouns - 1st Per. Plural |  |  |  | 0.54 |
| SANS11 - Inc. Latency |  | 0.63 | Sentiment - Valence |  |  | 0.44 | 0.68 |
|  |  |  | Sentiment - Arousal |  |  |  | -0.79 |
|  |  |  | Sentiment - Dominance |  |  |  | 0.76 |

### 3.2 Relationships between Social Cognition and Clinical Speech Ratings

Accounting for multiple comparisons, disorganized speech was significantly correlated with impairments in both emotion processing (r=- 0.56, p<0.001) and mentalization (r=-0.47, p<0.001). There was a trend for mentalization and underproductive speech (r=-0.27, p=0.06). Attribution bias was not significantly related to either clinical component. Figure 1A illustrates these relationships and effect sizes for individual clinical items.

**Figure 1.**
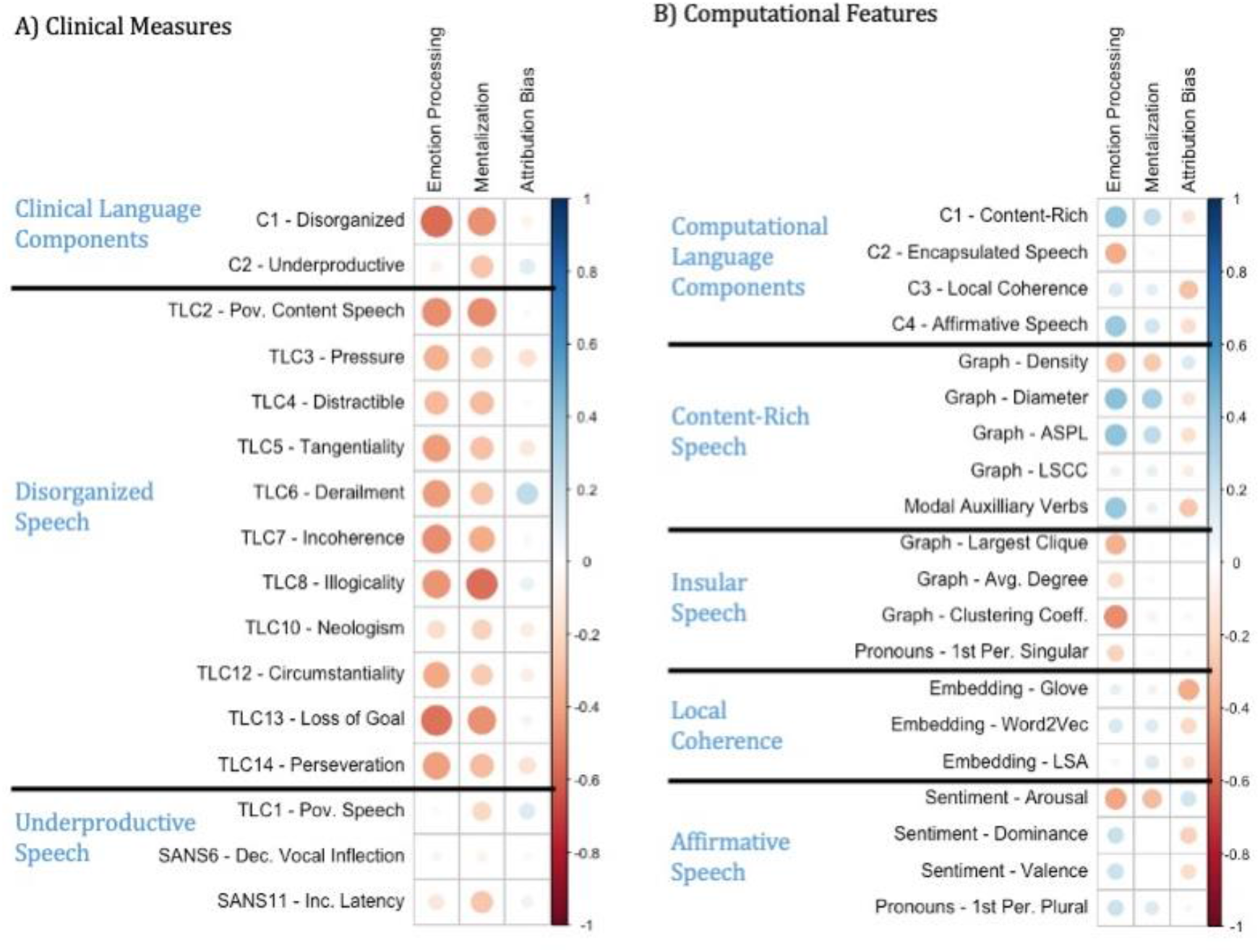
Correlations between Social Cognition and Measures of Speech Disturbance: Spearman correlation coefficients are illustrated for: A) Clinical Measures, including two principal components describing disorganized speech and underproductive speech; B) Computational Features, including four principal components describing content-rich speech, insular speech, local coherence, and affirmative speech.

Figure 2 shows scatterplots and linear best fits for social cognition and clinical speech components. We examined data collection method, age, sex, education level, overall psychosis symptoms and verbal ability as potentially confounding variables. However, disorganized speech continued to be significantly related to emotion processing (***β*** =-0.58 to -0.47) as well as mentalization (***β*** =-0.54 to -0.43), and underproductive speech continued to be significantly related to mentalization (***β*** =-0.39 to - 0.32).

**Figure 2.**
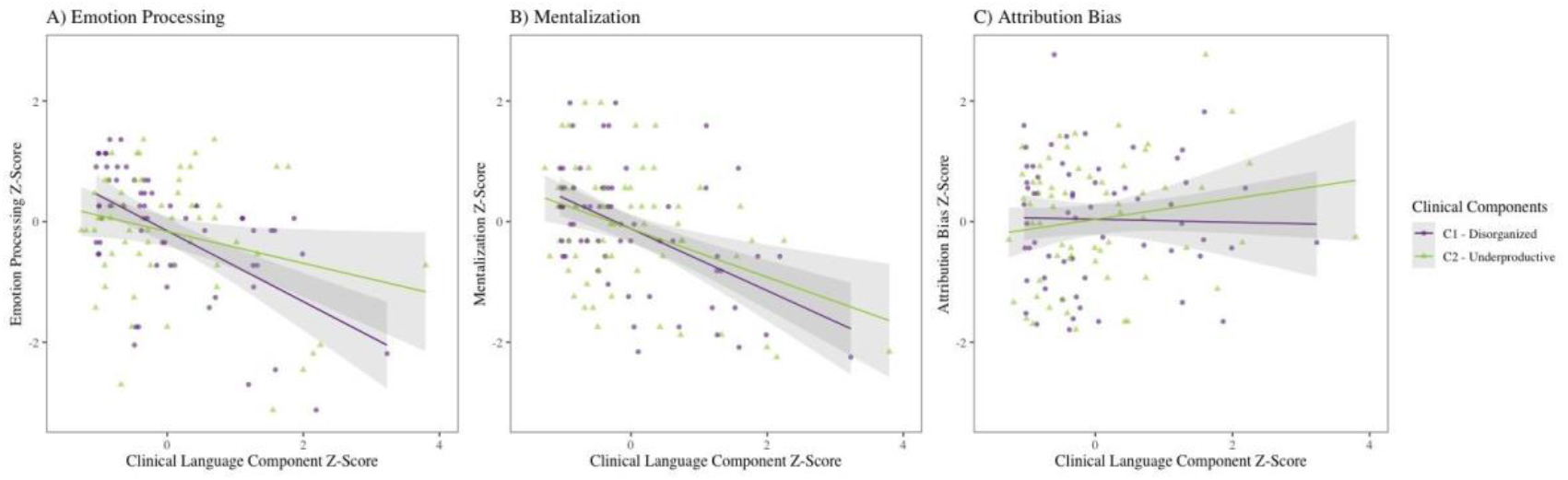
Associations between Clinical Language Components and Social Cognition: Scatterplots and linear best fits for: A) Emotion Processing, B) Mentalization, and C) Attribution Bias.

### 3.3 Relationships between Social Cognition and Computational Speech Features

Figure 1B illustrates correlations among social cognition domains, computational components and individual features. After adjusting for multiple comparisons, emotion processing ability was correlated with higher content-rich speech (r=0.41, p<0.05), lower insular speech (r=-0.37, p<0.05), and higher affirmative speech (r=0.38, p<0.05). Other notable trends were between mentalization and content-rich speech (r=0.27) as well as between attribution bias and local coherence (r=- 0.32); however, these relationships were not significant after accounting for the multiple comparisons.

Figure 3 shows scatterplots and linear best fits for social cognition and computational language components. Emotion processing was significantly predicted by insular speech even when accounting for all covariates (***β*** =-0.32 to - 0.27). However, the relationship between emotion processing and content-rich speech was no longer significant when accounting for education (***β*** =0.30, p=0.07) or verbal ability (***β*** =0.19, p=0.25); also, the relationship with affirmative speech was no longer significant when accounting for psychosis symptoms (***β*** = 0.23, p = 0.11) or data collection method (***β*** = 0.32, p = 0.13). Mentalization was significantly related to content-rich speech when accounting for data collection method, age, sex, psychosis symptoms, and verbal ability (***β*** = 0.29 to 0.35), but not when covarying for education level (***β*** =0.23, p=0.17). Attribution bias was related to local coherence when accounting for demographic variables and verbal ability (***β*** =-0.38 to -0.35) but trend-level when covarying for psychosis symptoms (***β*** =-0.26, p=0.07) and data collection method (***β*** =-0.32, p=0.06).

**Figure 3.**
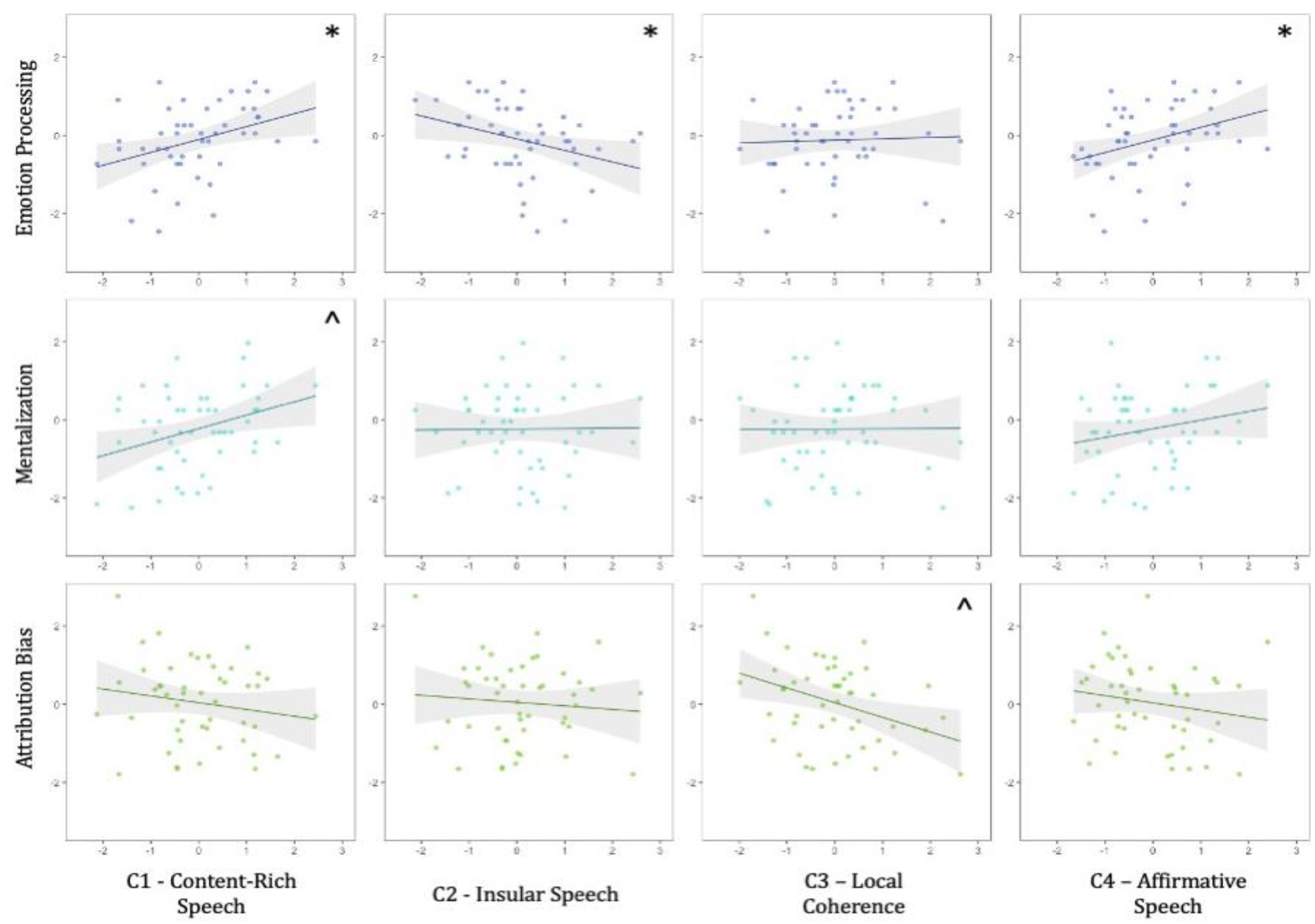
Association between Computational Language Components and Social Cognition: Z-scores for social cognition and computational component scores were compared. * Significant relationships. ^ Notable trends.

### 3.4 Longitudinal Exploratory Analyses

In the subsample of participants who returned for a followup assessment at 12 weeks (n=17), we explored whether changes in language measures predicted longitudinal changes in social cognition. Among clinical measures, improvement in mentalization was predicted by reductions in both disorganized (***β*** =-0.49, p<0.001) and underproductive speech (***β*** =-0.37, p<0.01). Improvement in emotion processing was predicted by reductions in disorganized speech (***β*** =-0.59, p<0.001), with a trend for underproductive speech (***β*** =-0.23, p=0.06). There were no significant longitudinal relationships among social cognitive domains and computational language features.

## 4. Discussion

In this study, we found strong evidence that emotion processing and mentalization are related to language disturbance in SSD, and particularly disorganized speech. On the other hand, attribution bias largely did not demonstrate consistent relationships with language measures. Using clinical ratings for disorganized and underproductive speech, we found that disorganization significantly predicted emotion processing and mentalization both within a single timepoint and in a longitudinal exploratory analysis. The cross-sectional relationships remained significant even when accounting for potential demographic, clinical, cognitive and methodological confounding variables. There was also evidence that underproductive speech relates to emotion processing and mentalization. However, there were no significant correlations between attribution bias and either clinical language component. Our results were consistent with those of the previous meta-analysis, which found consistent relationships between speech disturbance and both emotion processing and mentalization, but relatively sparse evidence for attribution bias (de Sousa et al., 2019). The effect sizes we report are somewhat larger, but in a similar moderate range.

Our findings were largely recapitulated using objective computational language features. Impairment in emotion processing was significantly predicted by insular speech, which is characterized by large quantities of speech on the same interconnected topics (in contrast to speech which explores multiple content areas). This relationship remained significant even when accounting for data collection method, demographic variables, psychosis severity, and verbal ability. Initially, emotion processing ability was also correlated with more content-rich speech and more affirmative speech. However, the correlation with content-rich speech may be explained by confounding relationships with education level and verbal ability, and the correlation with affirmative speech may be explained by a confounding relationship with psychosis severity. There was a notable trend between mentalization and content-rich speech, but this was not significant when accounting for education level. Also, there was a trend between higher attribution bias and reduced local coherence, but this may be explained by a confounding relationship with psychosis severity. Few other studies have directly evaluated social cognition and computational language features. However, our results are consistent with previous findings that social cognition and metacognition are associated with objective measures of syntactic complexity and cognitive complexity, respectively (Buck et al., 2015; Minor et al., 2019). Furthermore, psychosis in general has been associated with lower local coherence and negative valence (Birnbaum et al., 2020; Corcoran et al., 2018; Elvevåg et al., 2007); therefore, it is plausible that similar features may be associated with social cognitive impairment.

Overall, the results support our initial hypotheses and are consistent with the active inference model of discourse which posits that accurate updating of priors based on internal and external (including social) cues is required for receptive and productive language (Brown and Kuperberg, 2015; Palaniyappan and Venkatasubramanian, 2022). Emotion processing (or alternatively, simulation) and mentalization could therefore be considered crucial components of linguistic functioning. In contrast, attribution bias is not directly involved in language production, but might be a separate output of another active inference process. As expected, we found that emotion processing and mentalization showed fairly consistent relationships with clinical and computational language measures, while attribution bias was not strongly related to language, especially when accounting for overall psychosis severity. As an alternative to the active inference model, social cognition and language may also be connected via non-social neurocognition. That is, general cognitive impairments may cause both poor social cognitive performance as well as language disturbances. We did not directly assess all domains of non-social cognition, but we did find that verbal ability did not account for the relationship between disorganized speech and emotion processing or mentalization. Previous work also found that social cognition predicted additional variance in communication errors beyond a multidimensional assessment of neurocognition (Docherty et al., 2013). Moreover, if neurocognitive impairments were the driver of these relationships, we would have expected that social cognition would be more closely related to underproductive speech because of the close links between neurocognitive impairment and negative symptoms (which include alogia). Instead, we found that disorganized speech was more closely related to emotion processing and mentalization than underproductive speech. In another alternative, the universal human grammar describing cause and effect has been proposed as the basis for all human thought (Hinzen, 2014). Viewed through this framework, there might be primary disruptions in the grammar apparatus and semantic networks of individuals with SSD, which then cause impairments in social cognition. This explanation is not contradictory to the active inference model, as there could be a pathological feedback loop of language and social cognitive impairments.

On the whole, there were slightly larger effect sizes and more consistent findings for emotion processing than for mentalization. This is superficially surprising because mentalization seems more conceptually tied to the generation of a shared conversational framework. However, there are several potential explanations for this finding. The speech samples used for computational analyses were taken from tasks with little interaction between conversation partners. It may be that mentalization would be more closely related to language features from an interactive task (Achim et al., 2015). Also, mentalization is a less tangible ability than emotion processing, and therefore more difficult to measure objectively. The Hinting Task requires subjective judgements which may introduce noise and therefore weaken the results for mentalization.

There are direct clinical implications if we assume that there is a causal relationship between social cognition and language in SSD. Several effective social cognitive treatments have been developed, and some have been found to broadly improve outcomes (Horan and Green, 2019; Penn et al., 2007; Tang et al., 2020). Recent investigations also explore the role of targeted transcranial magnetic stimulation in improving social cognition (Oliver et al., 2021). If disruptions in active inference operations lead to psychosis symptoms, and if mentalization and emotion processing are critical for accurate feedback in social situations, then this further motivates the development of therapeutic strategies which target social cognition. Inversely, targeting linguistic functioning might improve social processing and functioning. It has been suggested that cognitive behavioral therapy (CBT) may improve psychosis symptoms by normalizing propositional meaning and deictic references—i.e., how we think about cause and effect and navigate shared assumptions (Zimmerer et al., 2017).

While suggestive, the current study neither provides proof for the active inference model of discourse nor establishes causal relationships between social cognition and language disturbance in SSD. In addition, our sample size was limited, particularly for the exploratory longitudinal analyses. There were several notable trends, including in the longitudinal analysis of computational features, which were not statistically significant but which were consistent with our overall findings. Further study in larger cohorts should be conducted to determine whether these were Type II errors. It is difficult to tease apart effects of data collection methods (in-person, virtual, or app-based) because different methods were used for different patient populations. Likely, the reported effects of data collection methods on our results are better explained by ascertainment differences. We relied on single measures for each social cognition domain. Convergent results from different tasks would provide further confidence for our findings. Similarly, studying the relationship between social cognition and receptive as well as productive language functioning would also provide further clarity.

Altogether, our results indicate that language disturbance in SSD is closely related to emotion processing and mentalization ability, but not attribution bias. This is particularly true for disorganized speech. Objective computational measures of speech also reflect these findings. Moreover, exploratory results suggest that longitudinal changes in social cognition are reflected by changes in speech measures. These findings are consistent with an active inference model of discourse, which could be a powerful framework for understanding psychosis pathology.

## Data Availability

All data produced in the present study are available upon reasonable request to the authors.

## Acknowledgements

We thank the participants for their contributions. We are also grateful to the clinicians and administrators at Zucker Hillside Hospital for their support of our work, including Drs. Michael Birnbaum, Anna Costakis, and Ema Saito. We thank Bill Simpson, Danielle De Souza, Jessica Robin, and Liam Kaufman of Winterlight Labs for their contributions to the ongoing collaborations.

## References

Achim, A.M., Achim, A., Fossard, M., 2017. Knowledge likely held by others affects speakers’ choices of referential expressions at different stages of discourse. Lang. Cogn. Neurosci. 32, 21–36. https://doi.org/10.1080/23273798.2016.1234059

Achim, Amelie M., Fossard, M., Couture, S., Achim, Andre, 2015. Adjustment of speaker’s referential expressions to an addressee’s likely knowledge and link with theory of mind abilities. Front. Psychol. 6. https://doi.org/10.3389/fpsyg.2015.00823

American Psychiatric Association, 2013. Diagnostic and statistical manual of mental disorders (DSM-5®).

Andreasen, N., 1979. Thought, language and communication disorders. Arch. Gen. Psychiatry 36, 1315–1321. https://doi.org/10.4324/9780203464199_chapter_4

Andreasen, N.C., 1986. Scale for the assessment of thought, language, and communication (TLC). Schizophr. Bull. 12, 473–482. https://doi.org/10.1093/schbul/12.3.473

Bedi, G., Carrillo, F., Cecchi, G.A., Slezak, D.F., Sigman, M., Mota, N.B., Ribeiro, S., Javitt, D.C., Copelli, M., Corcoran, C.M., 2015. Automated analysis of free speech predicts psychosis onset in high-risk youths. Npj Schizophr. 1. https://doi.org/10.1038/npjschz.2015.30

Benjamini, Y., Hochberg, Y., 1995. Controlling the False Discovery Rate: A Practical and Powerful Approach to Multiple Testing. J. R. Stat. Soc. Ser. B 57, 289–300.

Birnbaum, M.L., Norel, R., Meter, A.V., Ali, A.F., Arenare, E., Eyigoz, E., Agurto, C., Germano, N., Kane, J.M., Cecchi, G.A., 2020. Identifying signals associated with psychiatric illness utilizing language and images posted to Facebook. Npj Schizophr. 1–10. https://doi.org/10.1038/s41537-020-00125-0

Bowie, C.R., Gupta, M., Holshausen, K., 2011. Disconnected and underproductive speech in schizophrenia: Unique relationships across multiple indicators of social functioning. Schizophr. Res. 131, 152–156. https://doi.org/10.1016/j.schres.2011.04.014

Brown, M., Kuperberg, G.R., 2015. A Hierarchical Generative Framework of Language Processing: Linking Language Perception, Interpretation, and Production Abnormalities in Schizophrenia. Front. Hum. Neurosci. 9. https://doi.org/10.3389/fnhum.2015.00643

Buck, B., Iwanski, C., Healey, K.M., Green, M.F., Horan, W.P., Kern, R.S., Lee, J., Marder, S.R., Reise, S.P., Penn, D.L., 2017. Improving measurement of attributional style in schizophrenia; A psychometric evaluation of the Ambiguous Intentions Hostility Questionnaire (AIHQ). J. Psychiatr. Res. 89, 48–54. https://doi.org/10.1016/j.jpsychires.2017.01.004

Buck, B., Minor, K.S., Lysaker, P.H., 2015. Differential lexical correlates of social cognition and metacognition in schizophrenia; A study of spontaneously-generated life narratives. Compr. Psychiatry 58, 138–145. https://doi.org/10.1016/j.comppsych.2014.12.015

Combs, D.R., Penn, D.L., Wicher, M., Waldheter, E., 2007. The Ambiguous Intentions Hostility Questionnaire (AIHQ): A new measure for evaluating hostile social-cognitive biases in paranoia. Cognit. Neuropsychiatry 12, 128–143. https://doi.org/10.1080/13546800600787854

Corcoran, C.M., Carrillo, F., Fernández-Slezak, D., Bedi, G., Klim, C., Javitt, D.C., Bearden, C.E., Cecchi, G.A., 2018. Prediction of psychosis across protocols and risk cohorts using automated language analysis. World Psychiatry 17, 67–75. https://doi.org/10.1002/wps.20491

Corcoran, R., Mercer, G., Frith, C.D., 1995. Schizophrenia, symptomatology and social inference: Investigating “theory of mind” in people with schizophrenia. Schizophr. Res. 17, 5–13. https://doi.org/10.1016/0920-9964(95)00024-G

Couture, S.M., Penn, D.L., Roberts, D.L., 2006. The functional significance of social cognition in schizophrenia: A review. Schizophr. Bull. 32, 44–63. https://doi.org/10.1093/schbul/sbl029

de Sousa, P., Sellwood, W., Griffiths, M., Bentall, R.P., 2019. Disorganisation, thought disorder and socio-cognitive functioning in schizophrenia spectrum disorders. Br. J. Psychiatry 214, 103–112. https://doi.org/10.1192/bjp.2018.160

Docherty, N.M., McCleery, A., Divilbiss, M., Schumann, E.B., Moe, A., Shakeel, M.K., 2013. Effects of social cognitive impairment on speech disorder in schizophrenia. Schizophr. Bull. 39, 608–616. https://doi.org/10.1093/schbul/sbs039

Dwyer, K., David, A.S., McCarthy, R., McKenna, P., Peters, E., 2019. Linguistic alignment and theory of mind impairments in schizophrenia patients’ dialogic interactions. Psychol. Med. https://doi.org/10.1017/S0033291719002289

Elvevåg, B., Foltz, P.W., Weinberger, D.R., Goldberg, T.E., 2007. Quantifying incoherence in speech: An automated methodology and novel application to schizophrenia. Schizophr. Res. 93, 304–316. https://doi.org/10.1016/j.schres.2007.03.001

First, M.B., Gibbon, M., 2004. The structured clinical interview for DSM-IV axis I disorders (SCID-I) and the structured clinical interview for DSM-IV axis II disorders (SCID-II). Compr. Handb. Psychol. Assess. 2, 134–143.

Friston, K., Frith, C., 2015. A Duet for one. Conscious. Cogn. 36, 390–405. https://doi.org/10.1016/j.concog.2014.12.003

Green, M.F., Bearden, C.E., Cannon, T.D., Fiske, A.P., Hellemann, G.S., Horan, W.P., Kee, K., Kern, R.S., Lee, J., Sergi, M.J., Subotnik, K.L., Sugar, C.A., Ventura, J., Yee, C.M., Nuechterlein, K.H., 2012. Social cognition in schizophrenia, part 1: Performance across phase of illness. Schizophr. Bull. 38, 854–864. https://doi.org/10.1093/schbul/sbq171

Green, M.F., Horan, W.P., Lee, J., 2015. Social cognition in schizophrenia. Nat. Rev. Neurosci. 16, 620–631. https://doi.org/10.1038/nrn4005

Green, M.F., Penn, D.L., Bentall, R., Carpenter, W.T., Gaebel, W., Gur, R.C., Kring, A.M., Park, S., Silverstein, S.M., Heinssen, R., 2008. Social cognition in schizophrenia: an NIMH workshop on definitions, assessment, and research opportunities. Schizophr Bull 34, 1211–1220. https://doi.org/10.1093/schbul/sbm145

Hinzen, W., 2014. What is Un-Cartesian Linguistics. Biolinguistics 226–257.

Honnibal, M., Johnson, M., 2015. An Improved Non-monotonic Transition System for Dependency Parsing, in: Proceedings of the 2015 Conference on Empirical Methods in Natural Language Processing. Association for Computational Linguistics, pp. 1373–1378.

Horan, W.P., Green, M.F., 2019. Treatment of social cognition in schizophrenia: Current status and future directions. Schizophr. Res. 203, 3–11. https://doi.org/10.1016/j.schres.2017.07.013

Iter, D., Yoon, J., Jurafsky, D., 2018. Automatic Detection of Incoherent Speech for Diagnosing Schizophrenia. Proc 5th Workshop Comput. Linguist. Clin. Psychol. 136–146. https://doi.org/10.18653/v1/w18-0615

Kircher, T., Bröhl, H., Meier, F., Engelen, J., 2018. Formal thought disorders: from phenomenology to neurobiology. Lancet Psychiatry 5, 515–526. https://doi.org/10.1016/S2215-0366(18)30059-2

Kohler, C.G., Richard, J.A., Brensinger, C.M., Borgmann-Winter, K.E., Conroy, C.G., Moberg, P.J., Gur, R.C., Gur, R.E., Calkins, M.E., 2014. Facial emotion perception differs in young persons at genetic and clinical high-risk for psychosis. Psychiatry Res. 216, 206–212. https://doi.org/10.1016/j.psychres.2014.01.023

Kohler, C.G., Turner, T.H., Bilker, W.B., Brensinger, C.M., Siegel, S.J., Kanes, S.J., Gur, R.E., Gur, R.C., 2003. Facial emotion recognition in schizophrenia: Intensity effects and error pattern. Am. J. Psychiatry 160, 1768–1774. https://doi.org/10.1176/appi.ajp.160.10.1768

Krell, R., Tang, W., Hänsel, K., Sobolev, M., Cho, S., Tang, S.X., 2021. Lexical and Acoustic Correlates of Clinical Speech Disturbance in Schizophrenia. Workshop 35th AAAI Conf. Artif. Intell. 2021, 9.

Landauer, T.K., Foltz, P.W., Laham, D., 1998. An introduction to latent semantic analysis. Discourse Process. 25, 259–284. https://doi.org/10.1080/01638539809545028

Loper, E., Bird, S., 2002. NLTK: the Natural Language Toolkit, in: Proceedings of the ACL-02 Workshop on Effective Tools and Methodologies for Teaching Natural Language Processing and Computational Linguistics. Association for Computational Linguistics (ACL),

Morristown, NJ, USA, pp. 63–70. https://doi.org/10.3115/1118108.1118117

Mikolov, T., Sutskever, I., Chen, K., Corrado, G., Dean, J., 2013. Distributed Representations of Words and Phrases and their Compositionality. ArXiv13104546 Cs Stat.

Minor, K.S., Willits, J.A., Marggraf, M.P., Jones, M.N., Lysaker, P.H., 2019. Measuring disorganized speech in schizophrenia: Automated analysis explains variance in cognitive deficits beyond clinician-rated scales. Psychol. Med. 49, 440–448. https://doi.org/10.1017/S0033291718001046

Moore, T.M., Reise, S.P., Gur, R.E., Hakonarson, H., Gur, R.C., 2015. Psychometric properties of the penn computerized neurocognitive battery. Neuropsychology 29, 235–246. https://doi.org/10.1037/neu0000093

Morgan, C.J., Coleman, M.J., Ulgen, A., Boling, L., Cole, J.O., Johnson, F.V., Lerbinger, J., Bodkin, J.A., Holzman, P.S., Levy, D.L., 2017. Thought Disorder in Schizophrenia and Bipolar Disorder Probands, Their Relatives, and Nonpsychiatric Controls. Schizophr. Bull. 43, 523–535. https://doi.org/10.1093/schbul/sbx016

Mota, N.B., Copelli, M., Ribeiro, S., 2017. Thought disorder measured as random speech structure classifies negative symptoms and schizophrenia diagnosis 6 months in advance. Npj Schizophr. 3, 1–10. https://doi.org/10.1038/s41537-017-0019-3

Mota, N.B., Vasconcelos, N.A.P., Lemos, N., Pieretti, A.C., Kinouchi, O., Cecchi, G.A., Copelli, M., Ribeiro, S., 2012. Speech graphs provide a quantitative measure of thought disorder in psychosis. PLoS ONE 7, 1–9. https://doi.org/10.1371/journal.pone.0034928

Oliver, L.D., Haltigan, J.D., Gold, J.M., Foussias, G., DeRosse, P., Buchanan, R.W., Malhotra, A.K., Voineskos, A.N., 2019. Lower-And higher-level social cognitive factors across individuals with schizophrenia spectrum disorders and healthy controls: Relationship with neurocognition and functional outcome. Schizophr. Bull. 45, 629–638. https://doi.org/10.1093/schbul/sby114

Oliver, L.D., Hawco, C., Homan, P., Lee, J., Green, M.F., Gold, J.M., DeRosse, P., Argyelan, M., Malhotra, A.K., Buchanan, R.W., Voineskos, A.N., 2020. Social Cognitive Networks and Social Cognitive Performance Across Individuals With Schizophrenia Spectrum Disorders and Healthy Control Participants. Biol. Psychiatry Cogn. Neurosci. Neuroimaging S2451902220303566. https://doi.org/10.1016/j.bpsc.2020.11.014

Oliver, L.D., Hawco, C., Viviano, J.D., Voineskos, A.N., 2021. From the Group to the Individual in Schizophrenia Spectrum Disorders: Biomarkers of Social Cognitive Impairments and Therapeutic Translation. Biol. Psychiatry S0006322321015961. https://doi.org/10.1016/j.biopsych.2021.09.007

Overall, J.E., Gorham, D.R., 1962. The Brief Psychiatric Rating Scale. Psychol. Rep. 10, 799–812. https://doi.org/10.2466/pr0.1962.10.3.799

Palaniyappan, L., Mota, N.B., Oowise, S., Balain, V., Copelli, M., Ribeiro, S., Liddle, P.F., 2019. Speech structure links the neural and socio-behavioural correlates of psychotic disorders. Prog. Neuropsychopharmacol. Biol. Psychiatry 88, 112–120. https://doi.org/10.1016/j.pnpbp.2018.07.007

Palaniyappan, L., Venkatasubramanian, G., 2022. The Bayesian brain and cooperative communication in schizophrenia. J. Psychiatry Neurosci. 47, E48–E54. https://doi.org/10.1503/jpn.210231

Penn, D.L., Roberts, D.L., Combs, D., Sterne, A., 2007. The development of the social cognition and interaction training program for schizophrenia spectrum disorders. Psychiatr. Serv. 58, 449–451. https://doi.org/10.1176/ps.2007.58.4.449

Pennington, J., Socher, R., Manning, C.D., 2014. GloVe: Global Vectors for Word Representation. Proc. 2014 Conf. Empir. Methods Nat. Lang. Process. 1532–1543.

Pinheiro, J., Bates, D., DebRoy, S., Sarkar, D., R Core Team, 2022. nlme: Linear and Nonlinear Mixed Effects Models.

Pinkham, A.E., Harvey, P.D., Penn, D.L., 2018. Social Cognition Psychometric Evaluation: Results of the Final Validation Study. Schizophr. Bull. 44, 737–748. https://doi.org/10.1093/schbul/sbx117

R Core Team, 2016. R: A Language and Environment for Statistical Computing. Vienna, Austria.

Revelle, W., 2021. psych: Procedures for Psychological, Psychometric, and Personality Research. Northwestern University, Evanston, Illinois.

Schmidt, S.J., Mueller, D.R., Roder, V., 2011. Social cognition as a mediator variable between neurocognition and functional outcome in schizophrenia: Empirical review and new results by structural equation modeling. Schizophr. Bull. 37. https://doi.org/10.1093/schbul/sbr079

Snelbaker, A.J., Wilkinson, G.S., Robertson, G.J., Glutting, J.J., 2001. Wide Range Achievement Test 3 (wrat3), in: Understanding Psychological Assessment. Springer US, pp. 259–274. https://doi.org/10.1007/978-1-4615-1185-4_13

Solot, C.B., Moore, T.M., Crowley, T.B., Gerdes, M., Moss, E., McGinn, D.E., Emanuel, B.S., Zackai, E.H., Gallagher, S., Calkins, M.E., Ruparel, K., Gur, R.C., McDonald-McGinn, D.M., Gur, R.E., 2020. Early language measures associated with later psychosis features in 22q11.2 deletion syndrome. Am. J. Med. Genet. B Neuropsychiatr. Genet. 183, 392–400. https://doi.org/10.1002/ajmg.b.32812

Tan, E.J., Thomas, N., Rossell, S.L., 2014. Speech disturbances and quality of life in schizophrenia: Differential impacts on functioning and life satisfaction. Compr. Psychiatry 55, 693–698. https://doi.org/10.1016/j.comppsych.2013.10.016

Tang, S.X., Kriz, R., Cho, S., Park, S.J., Harowitz, J., Gur, R.E., Bhati, M.T., Wolf, D.H., Sedoc, J., Liberman, M., 2021. Natural Language Processing Methods are Sensitive to Sub-Clinical Linguistic Differences in Schizophrenia Spectrum Disorders. Npj Schizophr. 7, 25.

Tang, S.X., Moore, T.M., Calkins, M.E., Yi, J.J., Savitt, A., Kohler, C.G., Souders, M.C., Zackai, E.H., McDonald-McGinn, D.M., Emanuel, B.S., Gur, R.C., Gur, R.E., 2017. The Psychosis Spectrum in 22q11.2 Deletion Syndrome Is Comparable to That of Nondeleted Youths. Biol. Psychiatry 82. https://doi.org/10.1016/j.biopsych.2016.08.034

Tang, S.X., Seelaus, K.H., Moore, T.M., Taylor, J., Moog, C., O’Connor, D., Burkholder, M., Kohler, C.G., Grant, P.M., Eliash, D., Calkins, M.E., Gur, R.E., Gur, R.C., 2020. Theatre improvisation training to promote social cognition: A novel recovery-oriented intervention for youths at clinical risk for psychosis. Early Interv. Psychiatry 14, 163–171. https://doi.org/10.1111/eip.12834

Vasil, J., Badcock, P.B., Constant, A., Friston, K., Ramstead, M.J.D., 2020. A World Unto Itself: Human Communication as Active Inference. Front. Psychol. 11, 417. https://doi.org/10.3389/fpsyg.2020.00417

Warriner, A.B., Kuperman, V., Brysbaert, M., 2013. Norms of valence, arousal, and dominance for 13,915 English lemmas. Behav. Res. Methods 45, 1191–1207. https://doi.org/10.3758/s13428-012-0314-x

Wible, C.G., 2012. Schizophrenia as a Disorder of Social Communication. Schizophr. Res. Treat. 2012, 1–12. https://doi.org/10.1155/2012/920485

Yeo, I.-K., Johnson, R.A., 2000. A New Family of Power Transformations to Improve Normality or Symmetry. Biometrika 87, 954–959.

Zimmerer, V.C., Watson, S., Turkington, D., Ferrier, I.N., Hinzen, W., 2017. Deictic and Propositional Meaning—New Perspectives on Language in Schizophrenia. Front. Psychiatry 8. https://doi.org/10.3389/fpsyt.2017.00017

